# A Preliminary Study of Repetitive Transcranial Magnetic Stimulation for Cannabis Use Disorder and Its Effects on Concurrent Tobacco Use

**DOI:** 10.64898/2026.08.06.26359887

**Authors:** Masataka Wada, Nicole Petersen, Brendan Wong, Bohye Kim, Jane Paik Kim, Aimee McRae-Clark, Timothy Durazzo, Greg Sahlem

**Author notes:** Corresponding author: Masataka Wada, M.D., Ph.D., Department of Psychiatry and Behavioral Sciences, Stanford University, Palo Alto, USA, 401 Quarry Rd, Palo Alto, CA 94304.

## Abstract

**Objectives:** Tobacco and cannabis co-use is common, and reductions in one substance may theoretically lead to compensatory increases in the other. This secondary analysis examined whether cannabis cue-induced dorsolateral prefrontal cortex repetitive transcranial magnetic stimulation (DLPFC-rTMS) affects tobacco consumption among tobacco-using individuals with cannabis use disorder (CUD).

**Methods:** Data were analyzed from a randomized, sham-controlled trial of DLPFC-rTMS for CUD. Participants were treatment-seeking adults with moderate or severe CUD who reported baseline tobacco use. Active or sham 10-Hz DLPFC-rTMS was delivered during cannabis cue exposure over 10 treatment visits. Linear mixed-effects models examined group differences in weekly percentage change from baseline in tobacco consumption over treatment and follow-up, adjusting for baseline tobacco consumption. Additional models examined whether changes in cannabis use were associated with changes in tobacco use.

**Results:** Twenty participants were included, with 10 assigned to active rTMS and 10 to sham. Active rTMS was associated with a greater reduction in tobacco consumption than sham at 1-week post-treatment (t = −2.49, p = 0.015). Changes in cannabis use were not significantly associated with group differences in tobacco reduction. Estimated group differences did not indicate compensatory increases in tobacco use among participants with larger reductions in cannabis use.

**Conclusions:** Cannabis cue-induced DLPFC-rTMS was associated with a short-term reduction in tobacco consumption relative to sham among tobacco-using individuals with CUD. These preliminary findings suggest possible cross-substance effects of DLPFC-rTMS and did not indicate compensatory tobacco increases. Larger trials specifically designed for cannabis–tobacco co-use are warranted.

## Introduction

Cigarette smoking among U.S. adults has continued to decline, whereas cannabis use has increased over the past two decades ^1^. Tobacco use is common among individuals with CUD and is associated with greater cannabis-related clinical severity and poorer cannabis cessation outcomes ^2,3^. In addition, evidence suggests that cannabis use may hinder smoking cessation ^4^.

Repetitive transcranial magnetic stimulation applied to the dorsolateral prefrontal cortex (DLPFC-rTMS) has demonstrated efficacy as an intervention for tobacco use disorder (TUD) and evidence shows preliminary efficacy for the treatment of cannabis use disorder (CUD) ^5–7^. However, tobacco and cannabis use may function as substitutes for one another, raising concern that neuromodulation-induced reductions in one substance could be accompanied by compensatory increases in the other among individuals with dual use ^8,9^. Moreover, rTMS protocols for CUD are typically administered following cannabis-related cue exposure, whereas those for smoking cessation often use smoking-related cues. Despite the clinical relevance of cannabis-tobacco co-use, most neuromodulation studies have focused on a single substance-use outcome. As a result, it remains unclear whether rTMS protocols developed for one substance use disorder have beneficial, neutral, or potentially adverse effects on concurrent use of another substance. This question is particularly relevant for cue-based rTMS approaches, because the cue exposure used to engage addiction-related circuitry is typically substance-specific.

Therefore, this secondary analysis examined the effects of DLPFC-rTMS for CUD on tobacco consumption and its association with cannabis consumption among tobacco co-users.

## Methods

This secondary analysis used de-identified data from a previously published randomized controlled trial of DLPFC-rTMS for CUD conducted at Stanford University and the Medical University of South Carolina (NCT03144232). The trial was approved by both institutional review boards, and all participants provided written informed consent ^10^.

Participants were treatment-seeking adults aged 18–60 years who met DSM-5 criteria for moderate or severe CUD. Seventy-two participants were randomized. Participants received Active or Sham DLPFC-rTMS using the Beam-F3 method. Participants received 10-Hz rTMS over 10 visits, with two sessions per visit, across approximately five weeks. During each session, we administered stimulation while participants were exposed to cannabis-related cues. Each session consisted of 4,000 pulses delivered at a target intensity of 120% of resting motor threshold, with intensity adjusted for tolerability. Further details are reported in the original publication ^10^. We obtained clinical assessments at baseline, on the last day of each treatment week, and quantified weekly tobacco and cannabis use during a four-week post-treatment follow-up period.

To examine the longitudinal effects of rTMS for CUD on the change in tobacco use over the treatment period, we used a linear mixed-effects model. The weekly percent change from baseline of total tobacco consumption was specified as the outcome variable, and time, intervention, and their interaction were included as fixed effects. Baseline weekly tobacco consumption was also included as a covariate in the fixed-effects structure. The primary effect of interest was the interaction between time and intervention at each time point. To assess whether the effects of rTMS for CUD on changes in tobacco use were associated with concurrent changes in cannabis use, we added the percent change in cannabis consumption per week from baseline in the model and examined the three-way interaction between group, time, and the change in cannabis consumption. We additionally estimated the difference in the percent change in tobacco consumption at 1-week post-treatment between the groups at different levels of change in cannabis use sessions (25th, 50th, and 75th percentiles). No a priori power calculation was conducted because tobacco use was not a primary outcome of the parent trial.

## Results

We included a total of 20 participants in the analysis, excluding those who reported no tobacco use at baseline (**Table 1**). Ten participants were allocated to the Active group (mean age ± SD: 35.6 ± 9.54 years, male: 80%), and the remaining 10 were allocated to the Sham group (mean age ± SD: 33.3 ± 6.25 years, male: 70%). Baseline weekly tobacco consumption was 74.6 ± 59.6 in the Active group and 55.0 ± 52.0 in the Sham group. Baseline cannabis-use sessions per week were 22.0 ± 16.3 and 20.2 ± 13.2, respectively.

**Table 1:** Baseline demographic and substance-use characteristics of tobacco-using participants with cannabis use disorder.

| **Characteristic** | **Active rTMS (n = 10)** | **Sham rTMS (n = 10)** |
| --- | --- | --- |
| Age, years, mean ± SD | 35.6 ± 9.5 | 33.3 ± 6.2 |
| Male sex, n (%) | 8 (80%) | 7 (70%) |
| Baseline weekly tobacco consumption, mean ± SD | 74.6 ± 59.6 | 55.0 ± 52.0 |
| Baseline cannabis-use sessions per week, mean ± SD | 22.0 ± 16.3 | 20.2 ± 13.2 |

At 1-week post-treatment, the time-by-intervention interaction was statistically significant (β = −113.67 percentage points, t = −2.49, p = 0.015), indicating a greater reduction in tobacco consumption from baseline in the Active group than in the Sham group (Figure 1). Analysis including percent change in cannabis consumption from baseline did not show sufficient evidence of association between the change in cannabis use and the group difference in the change in tobacco consumption (β: 2.03, 95%CI: - 4.41 to 8.47).

**Figure 1.**
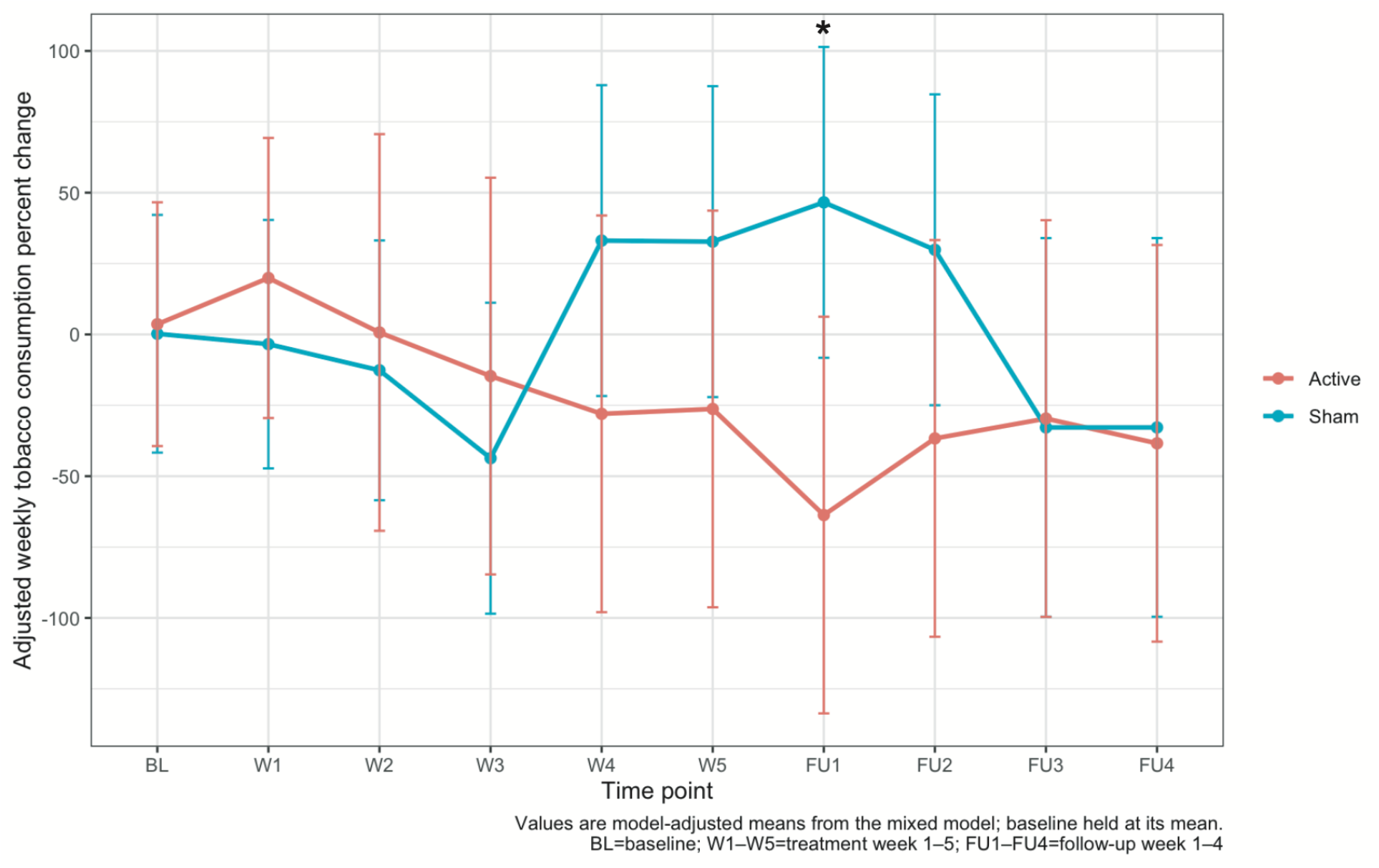
Weekly percent change from baseline in tobacco consumption over the course of rTMS treatment and during the four-week follow-up period. Points and lines represent model-adjusted means estimated from the mixed-effects model, with baseline tobacco consumption included as a covariate. Error bars indicate 95% confidence intervals.

However, the Active-Sham difference in weekly percent change from baseline of total tobacco consumption became smaller as cannabis use reduction became less pronounced. Because negative values of cannabis use change indicate reductions, the 25th percentile (−63.4) represented a larger reduction than the median (−52.6) or the 75th percentile (−28.9). The estimated group differences were −111.87 (95% CI: −214.9 to −8.80), −106.55 (95% CI: −205.0 to −8.12), and −94.80 (95% CI: −211.8 to 22.15), respectively. This suggests that our data do not support a compensatory increase in tobacco use following rTMS-induced reductions in cannabis.

## Discussion

In this exploratory secondary analysis, the Active and Sham groups differed in tobacco-consumption change at 1-week post-treatment, but the difference was not sustained at later assessments. Because stimulation was paired with cannabis cues, this finding may reflect cue-independent effects of DLPFC-rTMS or modulation of mechanisms shared across CUD and TUD. The transient effect suggests that protocol modifications may be needed to sustain reductions in tobacco use.

Bidzinski et al. (2022) reported a similar pattern in individuals with schizophrenia and CUD, indicating active DLPFC-rTMS was associated with a modest reduction in cigarette consumption, whereas the Sham group showed a transient increase ^11^. Together, these findings raise the possibility that rTMS may attenuate increases in tobacco use during cannabis reduction rather than directly produce a sustained reduction in tobacco use.

Point estimates did not indicate compensatory tobacco increases among participants with greater cannabis reductions. However, the wide confidence intervals and limited power for the three-way interaction preclude firm conclusions regarding cannabis-tobacco substitution. Thus, the findings should not be interpreted as evidence against substitution, but they support further investigation of cross-substance effects.

These findings may inform neuromodulation protocols for cannabis–tobacco co-use. Future trials should test whether tobacco-specific or combined cue exposure, booster sessions, or behavioral support improve and sustain effects on both substances.

Several limitations should be considered. Tobacco use was defined by current use rather than TUD diagnosis, tobacco cessation was not a treatment target, motivation to reduce tobacco use was not assessed, and substance use was self-reported. The small sample also limited statistical power, particularly for interaction effects, and produced imprecise estimates. Larger trials designed specifically for cannabis–tobacco co-use are needed.

## Conclusions

Cannabis-cue DLPFC-rTMS was associated with a between-group difference in tobacco-consumption change at 1-week post-treatment, but not with a sustained effect. These findings are hypothesis-generating and require confirmation in adequately powered trials.

## Author Contributions

Masataka Wada: Conceptualization, Methodology, Formal analysis, Writing - original draft. Nicole Petersen: Conceptualization, Methodology, Writing - review & editing. Brendan Wong: Investigation, Data curation, Writing – review & editing. Bohye Kim: Data curation, Writing - review & editing. Jane Paik Kim: Data curation, Writing – review & editing. Aimee McRae-Clark: Conceptualization, Investigation, Funding acquisition, Supervision, Writing - review & editing. Timothy Durazzo: Writing – review & editing. Greg Sahlem: Conceptualization, Methodology, Investigation, Funding acquisition, Supervision, Writing - review & editing. All authors reviewed and approved the final version of the manuscript and agree to be accountable for all aspects of the work.

## Data Availability Statement

The datasets analyzed during the current secondary analysis are available from the corresponding author upon reasonable request, subject to institutional review board approval, participant confidentiality requirements, and applicable data-sharing agreements. The data are not publicly available because public sharing is not permitted under the consent and privacy protections for the parent clinical trial.

